# An empirical cross-sectional analysis of the corrections in the New York Times’ COVID-19 coverage

**DOI:** 10.1101/2024.08.02.24311424

**Authors:** Alyson Haslam, Quiana Harshman, Vinay Prasad

**Author notes:** **Corresponding author:** Alyson Haslam, PhD, Department of Epidemiology and Biostatistics, UCSF Mission Bay Campus | Mission Hall: Global Health & Clinical Sciences Building | 550 16th St, 2nd Fl, San Francisco, CA 94158. **Disclosure:** V.P. receives research funding from Arnold Ventures through a grant made to UCSF, and royalties for books and writing from Johns Hopkins Press, MedPage, and the Free Press. He declares consultancy roles with UnitedHealthcare and OptumRX; He hosts the podcasts, Plenary Session, VPZD, Sensible Medicine, writes the newsletters, Sensible Medicine, the Drug Development Letter and VP’s Observations and Thoughts, and runs the YouTube channel Vinay Prasad MD MPH, which collectively earn revenue on the platforms: Patreon, YouTube and Substack. AH and QH report no disclosures. **Funding:** ORMJMP. The funder had no role in the design, data collection, data analysis, and reporting of this study.

## Abstract

**Background:** To examine the errors and corrections in the New York Times (NYT) and to assess if there is an imbalance towards overstating the pandemic severity, which may support more extreme restrictions or understating the severity.

**Methods:** We conducted a retrospective cross-sectional analysis of COVID-19 articles that had corrections made and reported in the NYT “corrections page”. We categorized authors in a NYT COVID-19 article as a NYT reporter, NYT other, or independent author. We calculated the number and type of corrections by author type (NYT reporter, NYT other, and independent author) and number and percentage of corrections indicating an over- or under-statement of the COVID-19 situation.

**Results:** There were 576 total corrections for the included 486 articles. Forty-three percent (n=245) corrections specifically pertained to COVID-19. Compared to corrections not pertaining to COVID-19, corrections pertaining to COVID-19 were less likely to be about spelling (0% vs 23.6%), locations (1.2% vs 16.3%), or title/degree (0% vs 10.6%), and more likely to be about a vaccine/vaccination (21.2% vs 0.3%), incidence/cases of conditions (12.2% vs 0.3%), or disease testing (7.8% vs 0.3%; p<0.001).

Compared to corrections not pertaining to COVID-19, corrections pertaining to COVID-19 were less likely to result in an equivocal tone (16.7% vs 88.8%), but they were more likely to both overstate (54.7% vs 8.5%) and understate (23.7% vs 2.4%) the situation in the original text (p<0.001). Ten reporters (of 346) accounted for 24% of the corrections. The reporter with the single most corrections accounted for 7% of the corrections.

**Conclusions:** Differential tone of the corrections suggests bias in the reporting of COVID-19 topics in a top news outlet. The reporting of unbiased information is a first step in addressing issues of misinformation in public health messaging.

## Background

The rapid speed at which the COVID-19 pandemic evolved resulted in a situation where the evaluation and validation of novel information was not able to keep up with the speed at which it was disseminated, thus resulting in the potential for errors. News reporters often must summarize or appraise evolving situations in limited time, with the potential risk of incorrect information.

It’s been estimated that about 11% of newspaper articles have errors that are brought to the attention of the publisher, [1] yet many of these errors are not corrected -with an estimated 2% of articles with errors receiving corrections, many of which are “low-impact”.[2]

Historically, journalists have been viewed as impartial distributors of information, but in a recent Pew survey, journalists are less likely to agree with impartiality in journalism than the general public, and people who lean towards a liberal political view are also less likely to agree that journalists should be impartial.[3] Even with these beliefs in impartiality, 60% of people in the US view news outlets as being politically biased.[4]

When errors are large or unidirectional, they may constitute misinformation. The spread of misinformation in addition to correct information during the pandemic has been described by the World Health Organization as an “infodemic”, and the spread of misinformation is believed to have resulted in the hospitalization of thousands and death of hundreds during the first few months of the pandemic.[5] Many have attributed the spread of misinformation largely to social media routes, whereas ascribing the reinforcement of public health recommendations to primarily news media.[6]

Indeed, as purveyors of trusted public health information, traditional news media sources have been described as “key” players in combating the misinformation epidemic.[7] Reasons for this include the perception of journalistic integrity and credibility, and having strict editorial standards,[8] which can lead to an environment where there is coordinated effort between government and local news stations in disseminating public health information.[9]

Because of the trusted position of the traditional media, there lies the responsibility to adhere to high-quality, truthful, and transparent reporting. Moreover, there is an obligation to be unbiased when reporting on topics, especially those perceived as polarizing or where there is equipoise. While the majority of information published in traditional media sources is correct, there are occasional mistakes that are made, and these are often corrected. We assume a non-partisan media source will have errors reported non-differentially – meaning that some errors are overstatements, some are understatements, and some do not result in a notable change in meaning.

It is from this perspective that we sought to examine the errors and corrections in NYT and to assess if there is an imbalance towards overstating the pandemic severity which may support more extreme restrictions or understating the severity. As a comparator group, we sampled corrections during the same time period that did not specifically pertain to COVID-19 and assessed whether they over- or under-stated the relevant or target problem.

## Methods

To assess the direction of corrected reporting, we searched the New York Times (NYT) because it was the most circulated news source in the United States, as per both digital and print subscriptions.[10] We searched the NYT website for articles that had had corrections made. We searched for articles by including the word “COVID” in the search bar of the Corrections page[11] on February 21, 2024.

We included articles that specifically discussed the topic of COVID-19 – namely those reporting on epidemiology, vaccines, masks, policy, funding, and/or government reactions to COVID-19. We excluded articles that reported on a singular death from COVID, events that occurred during the COVID-19 pandemic, but were not specifically related to COVID-19, epidemics in general, and governmental agency credibility. We also excluded articles not in English.

From included articles, we abstracted information on the author names of the article, the date the article was originally published, the correction information, the date of correction, and how many corrections were made. We classified the correction as a topic type (e.g., deaths, cases, vaccine or mask policy, spelling, date, etc.). We also classified the corrections as to whether they pertained specifically to COVID-19 or not and whether the error pertained to a number or statistic. We calculated the time interval between the date when the article was originally published and the date when the correction was made. If corrections were made on multiple days, we used the date of first correction. We abstracted data on author title/affiliation. If the writer was from the NYT, we classified them as a reporter, editor, columnist, correspondent, visual report/editor, bureau chief, opinion reporter/editor, or other (food critic, fashion editor, fellow). If the author was not one of these (e.g., academic professor, writer for other journal/newspaper/book), we classified it as independent. If more than 5 journalists contributed to an article, we classified these as being written by a single group (e.g., NYT), and we did not include them in the author analysis since it would be hard to determine who was responsible for the correction.

From the correction information, we classified it as having originally overstated the information, understated the information, or had no effect. If there wasn’t enough information to determine the original statement, we coded it as being unknown. In general, we classified statements originally reporting a more dire or more extreme situation as being overstatements. This included numbers that were higher than corrected numbers, in the instances of hospitalization, cases, or deaths, or lower numbers for vaccination status. Conceptually, more closures, later re-opening dates, or more resources invested into fighting COVID-19 in the original reporting were considered overstatements. The direction of the correction was coded by two blinded reviewers (AH and QH), and discordant coding was discussed for consensus.

For authors with one or more corrections, we searched to see how many total COVID-19 articles listed them as an author or coauthor between the dates of 02/01/2020 – 02/21/2024. This was done by searching the author’s name (in quotes) and the term “covid” in the general search bar, filtered by the specified dates. We then calculated a rate of corrections for each of them.

### Statistical analysis

We calculated descriptive statistics for the characteristics of the articles and corrections. We used Cohen’s Kappa to determine the level of agreement in the coding of the direction of the correction. Values between 0.41 and 0.60 indicated moderate agreement, 0.61 to 0.80 indicated substantial agreement, and 0.81 to 0.99 indicated almost perfect agreement.[12] We used a Chi-square test to determine differences in characteristics between COVID-19 corrections and non-COVID-19 corrections. We also tested differences in characteristics between author role (NYT reporter, NYT other, and independent author) using a Chi-square test. All analyses were done in Excel (Microsoft Corporation) and R statistical software (R Project for Statistical Computing), version 4.2.1.

In accordance with 45 CFR §46.102(f), this study was not submitted for University of California, San Francisco institutional review board approval because it involved publicly available data and did not involve individual patient data.

## Results

We found 2393 articles with corrections and having the term “COVID” in them, of which 486 reported specifically on COVID or a COVID-related topic. Three articles were written by the “editorial board” so the author number could not be determined, and one article did not report any information about the correction – only that there was a correction made.

There were 576 total corrections for the included 486 articles (Table). Each article had a median of one author (range: 1-56) and one error (range: 1-9). One article did not give specifics on the correction, only that there was a correction made. Corrections made by NYT reporters comprised 47.6% (n=274) of all corrections, followed by columnists (10.6%, n-61) and independent authors (10.4%, n=60). Forty-three percent (n=245) corrections specifically pertained to COVID-19. The most common types of corrections pertained to spelling (n=78; 13.5%), location (n=57; 9.9%), date/time (n=53; 9.2%), and vaccine/vaccination information (n=53; 9.2%). Overall, 25% (n=144) pertained to a statistical number. The median number of days from when the article was first published and when the correction was made was 1 day (range: 0-91) Most corrections were equivocal in their tone (n=408, 59.1%), while 27.7% (n=191) were overstated in the original text, 11.3% (n=78) were understated in the original text, and 1.9% (n=13) of corrections did not provide enough information to determine the direction of the correction.

**Table.**
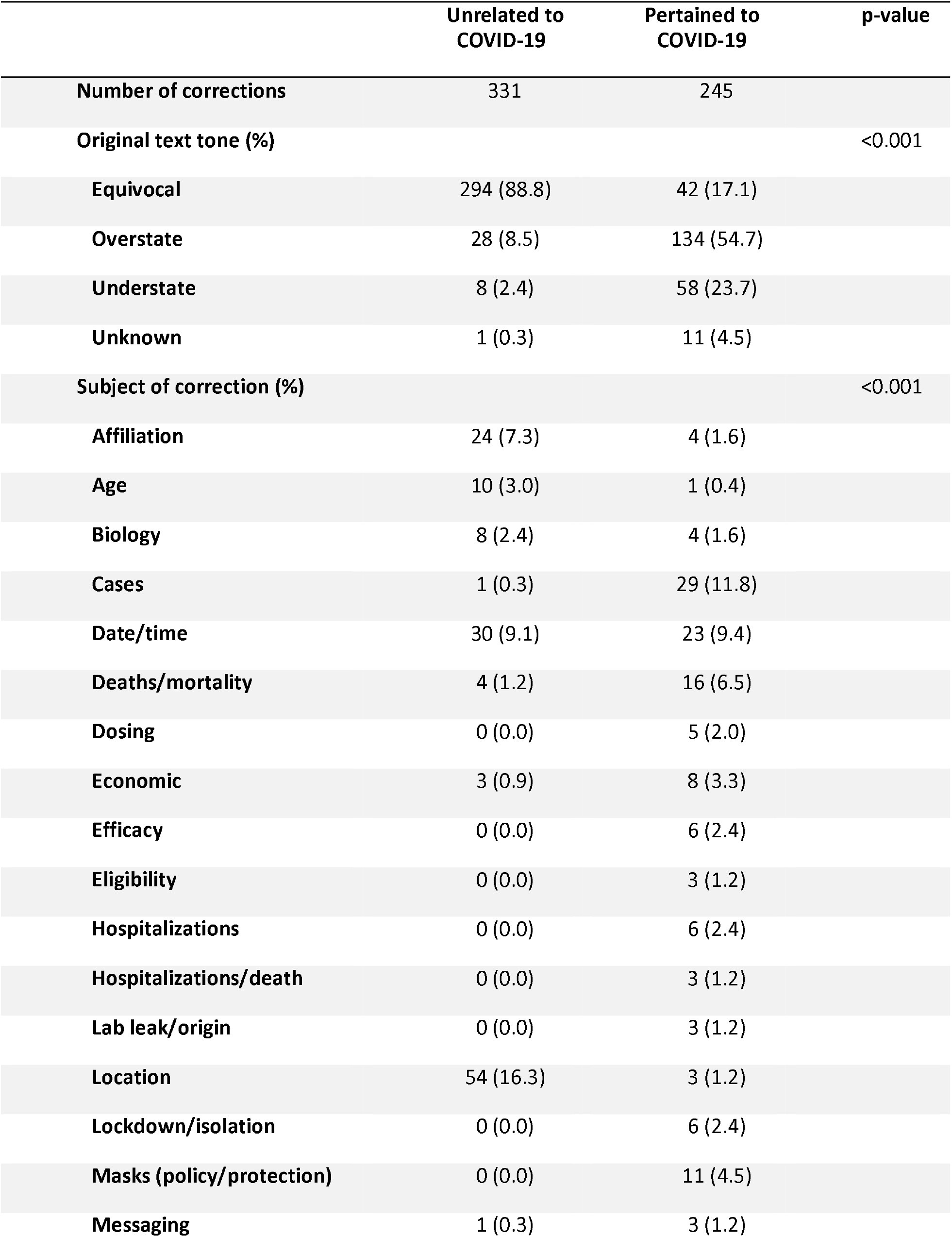
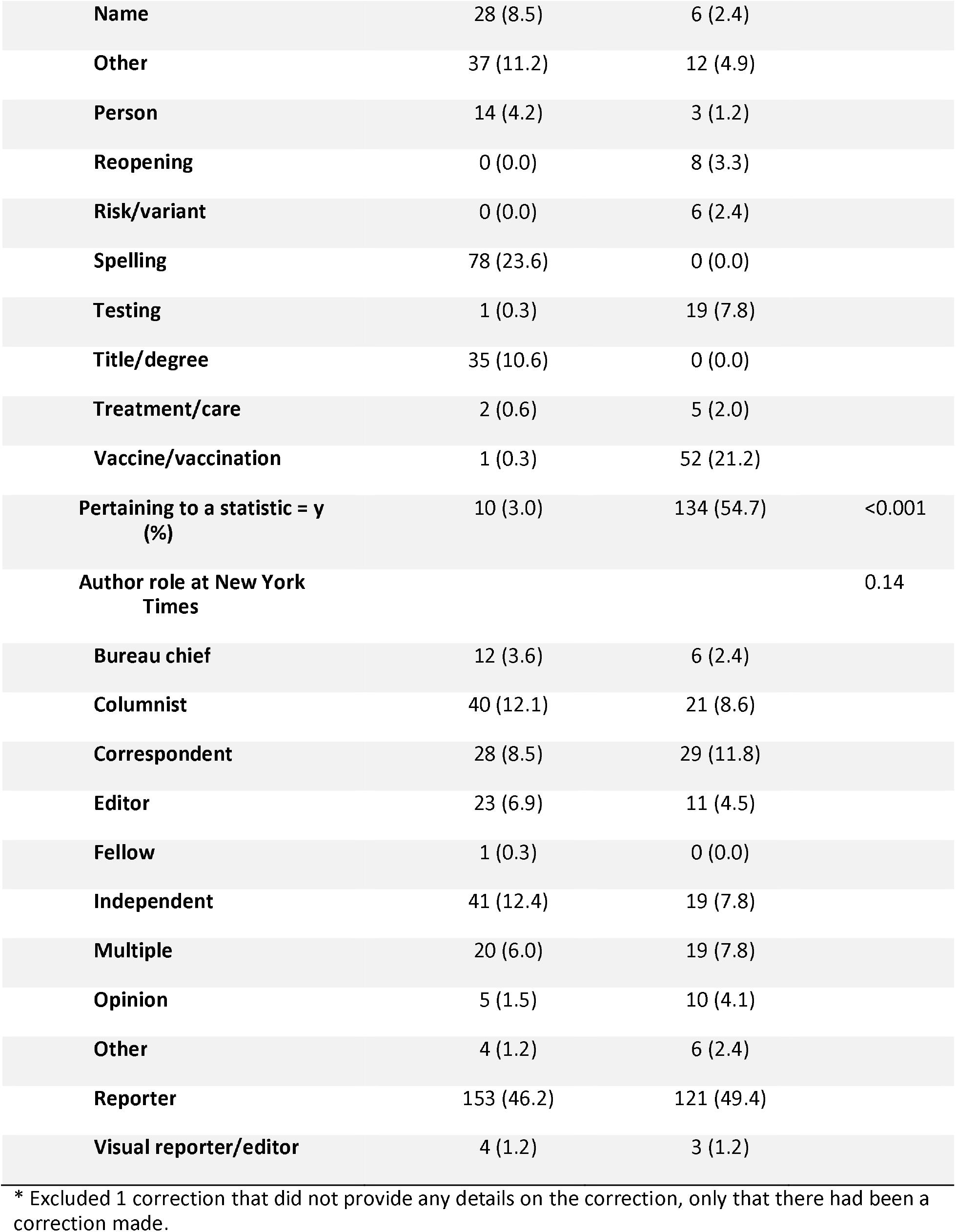
Characteristics of corrections for COVID-19 NYT articles, stratified by their role as an author*.

Compared to corrections not pertaining to COVID-19, corrections pertaining to COVID-19 were less likely to be about spelling (0% vs 23.6%), locations (1.2% vs 16.3%), or title/degree (0% vs 10.6%), and more likely to be about a vaccine/vaccination (21.2% vs 0.3%), incidence/cases of conditions (12.2% vs 0.3%), or disease testing (7.8% vs 0.3%; p<0.001).

Compared to corrections not pertaining to COVID-19, corrections pertaining to COVID-19 were less likely to result in an equivocal tone (16.7% vs 88.8%), but they were more likely to both overstate (54.7% vs 8.5%) and understate (23.7% vs 2.4%) the situation in the original text (p<0.001).

The kappa statistic for determining the level of agreement in the direction of the correction was 0.61 (p<0.001), indicating substantial agreement between raters.

There were 349 unique authors, if not considering unspecified authors or papers with more than 5 authors (e.g., “editorial board”, “NYT”, “The Daily”, etc). The median correction by each author was one (range: 1-55). Thirty-three percent of corrections made by NYT reporters had the original information overstated, whereas it was 26.0% for other NYT authors and 16.7% for independent authors (p=0.07; Table and Figure 1). Figure 2 shows the distribution of the number of overstated and understated corrections for authors. The median percentage of COVID-19 corrections per author was 2% (range: 0.3% to 100%), and the median percentages of overstatements and understatements were 0% (range: 0% to 100%) and 0% (range: 0% to 50%). Ten reporters (of 346) accounted for 24% of the corrections. The reporter with the single most corrections accounted for 7%. Figure 3 shows the distribution of the percentage of COVID-19 corrections indicating an overstatement and understatement of the original information for NYT reporters with at least 50 NYT COVID-19 articles.

**Figure 1.**
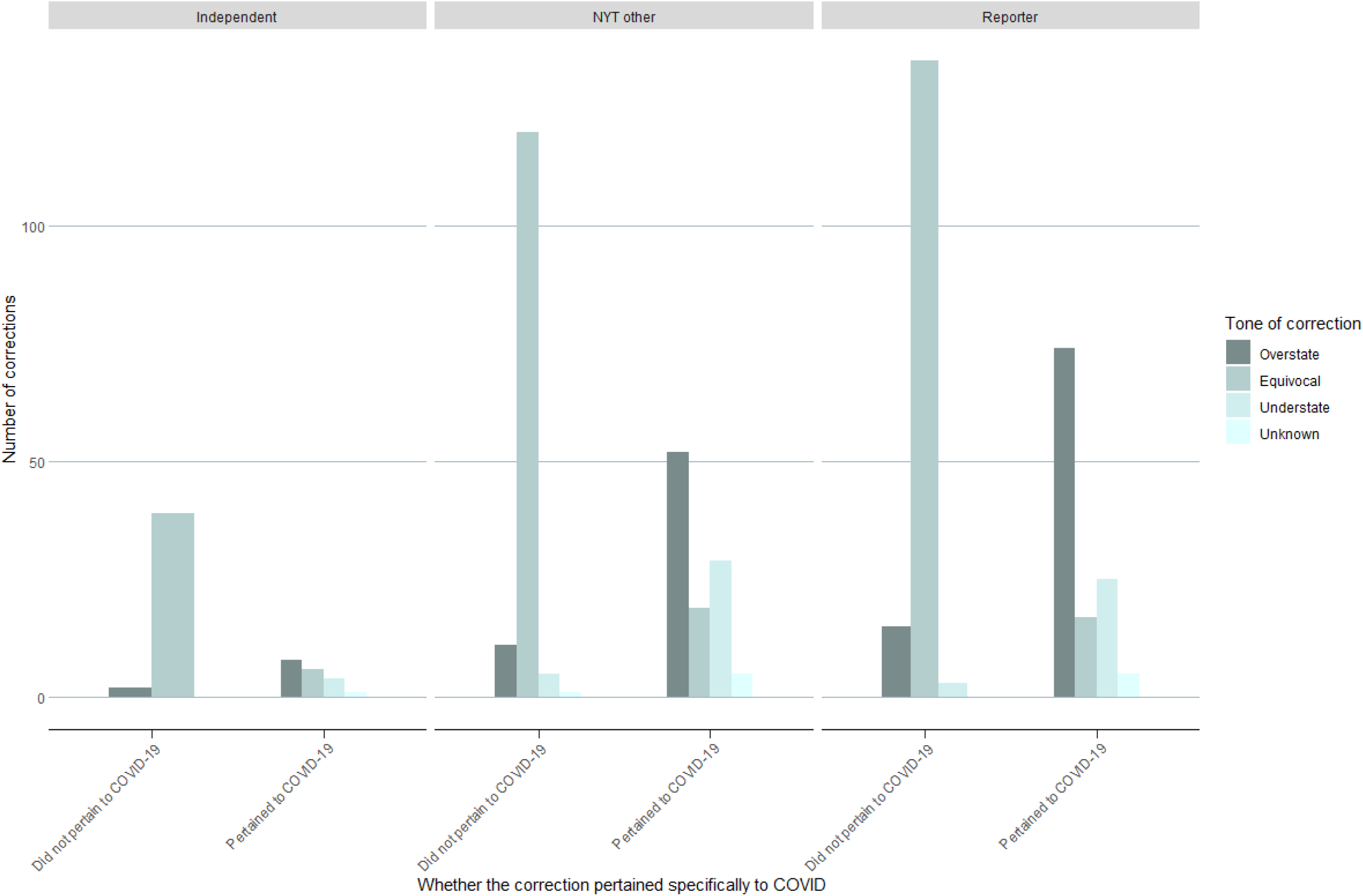
Number of New York Times corrections where the original text over or understated the COVID-19 situation, by whether the correction was specific to COVID-19 or not.

**Figure 2.**
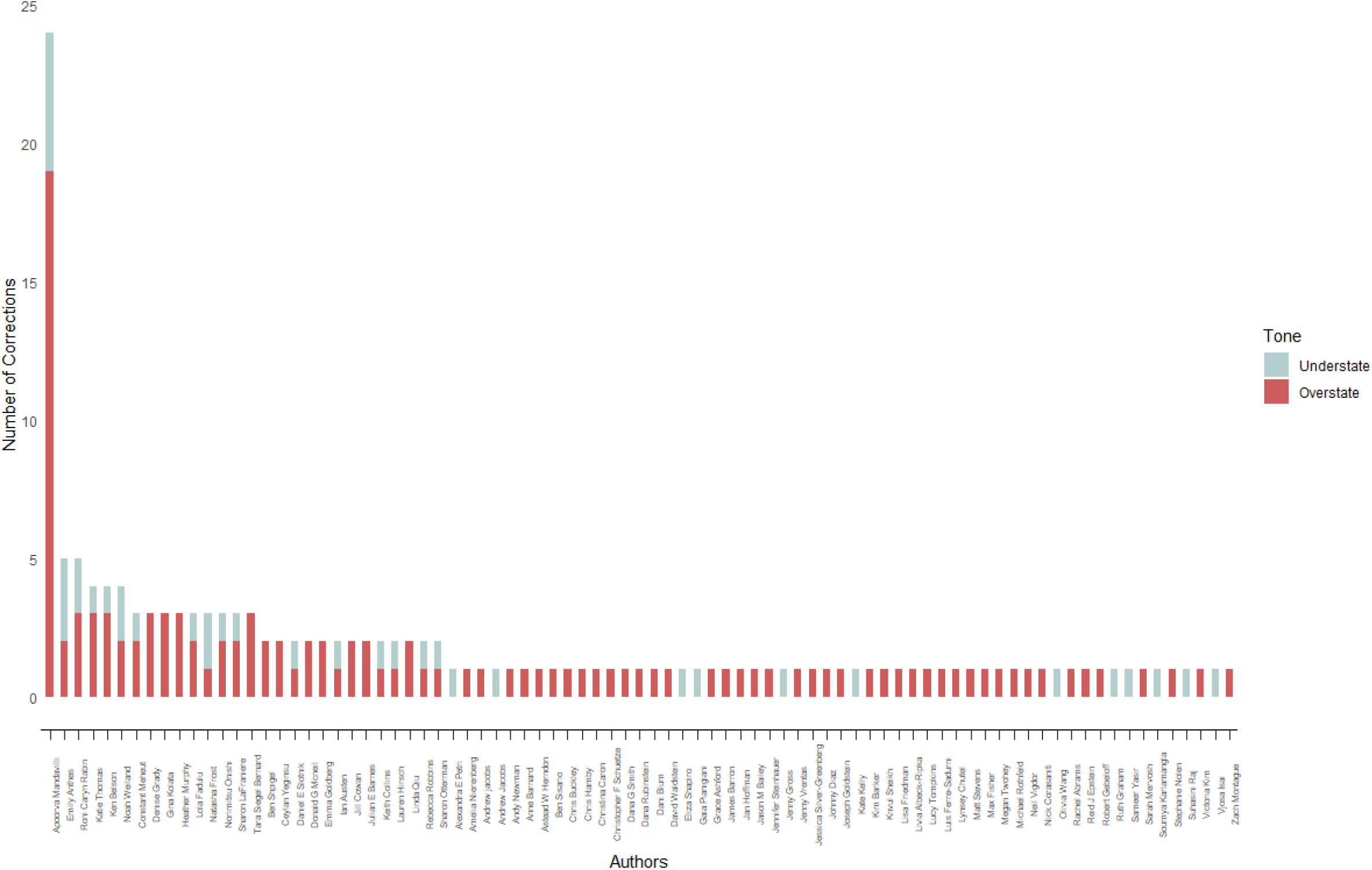
Number of corrections for each New York Times reporter on a COVID-19 article, including only those that were over or understatements in the original text.

**Figure 3.**
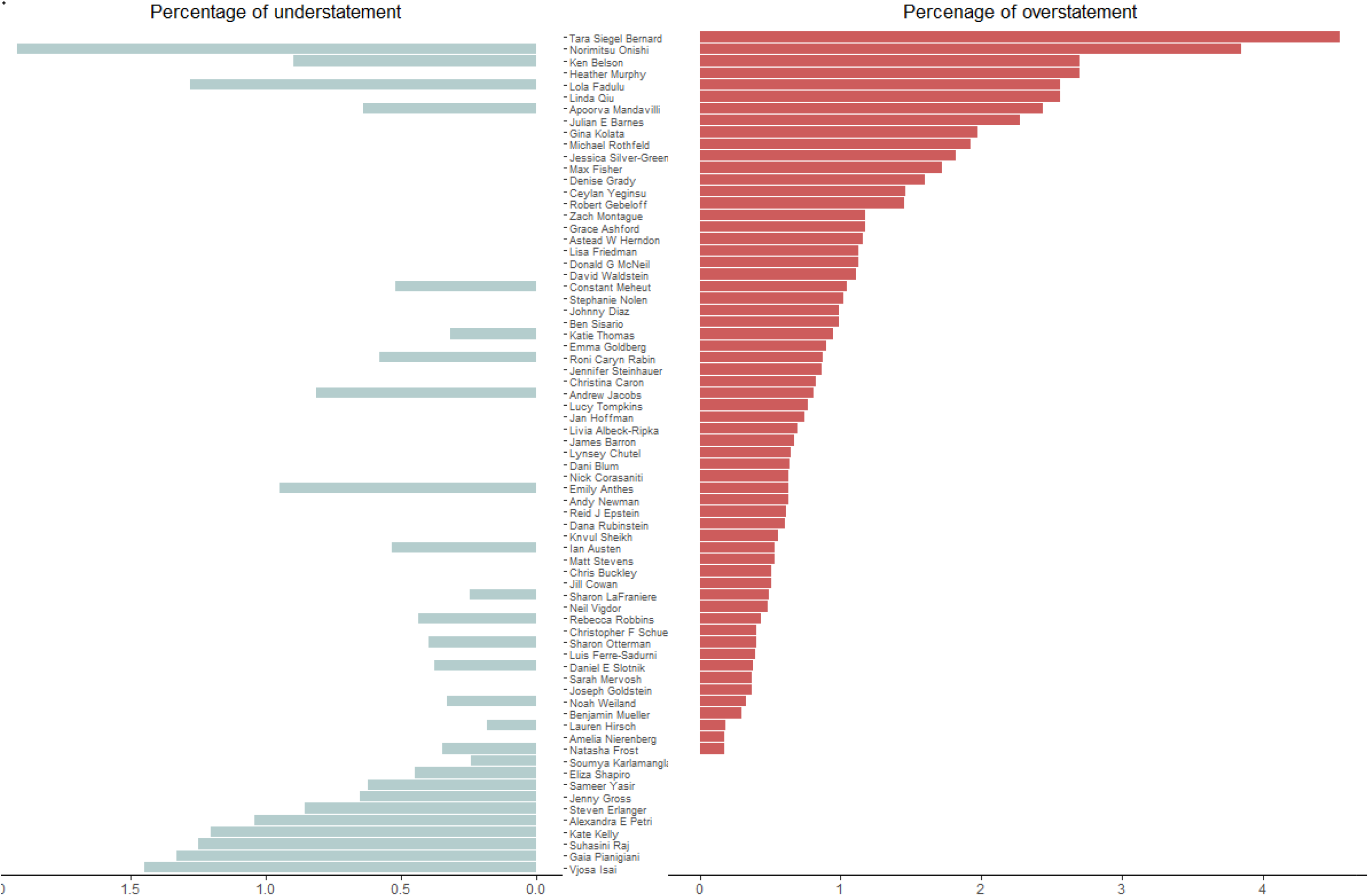
Percentage of overstatements and understatements for New York Times reporters (>50 articles) with corrections in New York Times COVID-19 articles.

## Discussion

We found that corrections made by NYT authors often led to meaningful differences in how information was portrayed, particularly when corrections pertained to COVID-19, where the original information more often overstated the COVID-19 pandemic and gave the impression that the situation was more dire than it actually was.

Overall, the majority of all corrections led to no meaningful difference in how the information was interpreted, unless the correction was about COVID-19. In these instances, the majority of corrections indicated a more exaggerated tone or more dire situation in the original publication. This is noteworthy considering that much of the research and discussion on misinformation during the COVID-19 pandemic has centered around social media’s role and not mainstream media’s role.[13] Conversely, mainstream media has been viewed as minimally a mediator of truth and more generously as a guardian of truth.[14, 15] Prior to the COVID-19 pandemic, it was estimated that only a part of fake news was disseminated through social media,[16] although trust even in the mainstream media had been declining.[15]

While most COVID-19 corrections indicated an overstatement of the originally reported information, another notable finding is that there was heterogeneity in whether authors routinely under or overstated the original information. Balanced reporting should lead to non-differential differences in the percentages of overstatements and understatements. We found that while a few authors had a similar percentage of both overstatements and understatements, most corrections by authors indicated a bias in one direction or another, and of those, most indicated an overstatement of the original information. Moreover, almost one-quarter of the corrections were made by fewer than 3% of all authors.

When corrections were made, the time between reporting and the information being corrected was short (median of one day). While it is understood that mistakes are made in reporting, credible, upstanding sources will make corrections quickly, although even quickly made corrections can lead to public distrust.[17] Additionally, transparent journalism should also include information on the correction. While most corrections contained information about both the correction and the original reporting, 2% of articles did not, and one did not provide any information about the correction, only the date that a correction was made.

In 2017, the NYT revised their editing process to a more “streamlined” process. As a result, the number of corrections decreased slightly, but a higher number of corrections were made in the front section, where readers are more likely to get information on world, national, and political news.[18] It is unknown how this process could have affected the corrections made in regards to reporting on the COVID pandemic, but likely the front section is where many COVID-19 stories would have been reported. The errors in these sections could have had more influence on public perception than errors contained in other sections.

### Limitations

First, our results may not be generalizable to all media sources or newspapers. We only selected one news source because it had the largest number of subscribers. But because it has the most number of subscribers, it would have more influence on influencing readers’ opinions. Because of the way that searches are done on the NYT, we had to use a different search strategy for identifying the denominator, but because the same method for determining the number of author papers was used for all authors, this should lead to non-differential bias in determining the percentage of corrections. And, while this may affect the exact rate of corrections, our focus was to show comparative trends in rates. Finally, because some studies had more than one author, and we were not able to determine which author actually made the error, the corrections from these articles were attributed to each author.

## Conclusion

We examined both COVID-19 specific and COVID-10 non-specific corrections in NYT articles covering COVID-19 topics. When compared to non-COVID-19 corrections, we found COVID-19 corrections were six times more likely to overstate the magnitude of the problem. In other words, when reporters erred, it was towards engendering greater fear and panic. We also found that individual reporters had different patterns of error. Eleven percent tended to underemphasize the pandemic, while 28% tended to overemphasize it before it was corrected. Less than 3% of authors were responsible for almost one-quarter of all corrections, and the reporter with the single most corrections accounted for 7% of all corrections. Our results suggest that corrections may plague some reporters more than others, warranting a more careful review of information prior to disseminating to the public.

## Data Availability

Data are available upon request to the author.

## Authorship statement

AH and VP conceptualized study design; AH abstracted data; AH and QH coded data; AH conducted statistical analysis and wrote first draft; QH and VP edited final draft.

